# Characterization and Resistance to Mutation of a Single-Channel Multiplex PCR Assay for SARS-CoV-2

**DOI:** 10.1101/2023.04.27.23289205

**Authors:** Amy L. Pednault, Brian M. Swalla

## Abstract

Throughout the COVID-19 pandemic, wastewater surveillance has been used worldwide to provide valuable public health data. RT-qPCR is frequently used as a quantitative methodology for wastewater surveillance but is susceptible to mutations in target regions. These mutations may lead to misinterpretation of surveillance data; a drop in signal could be concluded to be a result of lower viral load, when in fact it is caused by reduced detection efficiency. We describe a novel approach to mitigating the impacts of such mutations: monitoring the cumulative signal from two targets (N1 and N2) via independent amplification reactions using identically labeled probes; a “single-channel multiplex” approach. Using the IDEXX Water SARS-CoV-2 RT-qPCR test, we demonstrate equivalent intra-assay repeatability and quantitative results from the combined N1N2 test when compared to individual N1 and N2 assays. Furthermore, we show that while mutations in B.1.1.529, BA.5.2, and BA.5.2.1 significantly impact the performance of the N1 assay, the impact on the N1N2 assay was negligible, and nearly within acceptable margin of error for technical replicates. These findings demonstrate that a single-channel multiplex approach can be used to improve the robustness of wastewater surveillance and minimize the risk of future mutations leading to unreliable public health data.

## Introduction

Wastewater surveillance for severe acute respiratory syndrome coronavirus 2 (SARS-CoV-2) has provided valuable information to public health officials during the COVID-19 pandemic (Hopkins et al., 2023; Kirby et al., 2021), with programs implemented around the world (Agrawal et al., 2021; Arora et al., 2020; Prado et al., 2021). Notable examples include the National Wastewater Surveillance System (Kirby et al., 2021), as well as several national systems resulting from a European Commission recommendation issued in October of 2021 (European Commission, 2021). Surveillance has shown that trends in community infection dynamics correlate with trends in viral copies of SARS-CoV-2 in wastewater (Peccia et al., 2020). Wastewater surveillance can therefore provide valuable, independent information on infection trends via a single, non-invasive sampling event representing an entire community.

Quantitative reverse-transcription polymerase chain reaction (RT-qPCR) is commonly used for wastewater surveillance (Aw & Rose, 2012; Farkas et al., 2020). To be reliable, RT-qPCR requires appropriately designed primers and probe(s) that provide specific amplification and detection. When the target region displays sequence heterogeneity, additional design considerations are required to ensure all relevant sequences are detected. Various mutations in SARS-CoV-2 have emerged during the pandemic, and occasionally these mutations have impacted the target annealing regions for RT-qPCR tests.

The existence of an emerging mutation affecting a RT-qPCR test may go unrecognized until sequence data has been collected and analyzed. Additional time may be required to understand whether a new mutation is growing in prevalence and associated as a characteristic feature in a significant new variant. This delay, coupled with the need for prompt interpretation and response to wastewater surveillance trending data, creates a time during which responsive actions are taken before the significance of a new mutation is understood. This could lead to misinterpretation of recent trends; for example, it may be concluded that an apparent drop in viral detection indicates lower viral load in the wastewater, when in fact the lower viral measurements were caused by reduced RT-qPCR detection efficacy.

To mitigate this risk, it is important to implement primary surveillance methods for trending of SARS-CoV-2 in wastewater that are robust to the impacts of mutation. One widely used approach is to monitor multiple regions within the target genome. This approach provided the first evidence of an emerging mutation in the B.1.1.529 variant where failure of a specific PCR design targeting the S gene occurred while detection of other targets continued unaffected (Kidd et al., 2021). Although monitoring of multiple targets can be effective, there are limitations to this approach. First, the impact of a mutation on RT-qPCR can vary dramatically. For example, a deletion is more likely to cause a severe and observable assay defect compared to a single nucleotide substitution that may have little or no effect. Second, mutations with modest effects may remain undetected due to larger sources of variation in upstream sample handling and processing steps. Third, due to the increased complexity and variance of monitoring multiple targets, it can be challenging to determine when the convergence or divergence in results between targets may respectively indicate the absence or presence of a significant mutation. Monitoring multiple regions within the target genome, therefore, can cause lower confidence in results when a new variant is becoming more prevalent, but before adequate sequencing is performed. Finally, conducting multiple assays adds time, cost, and complexity to the laboratory workflow.

A different approach has been described in which detection of highly variable targets was improved by using identically labelled hydrolysis probes with a single primer pair (Nagy et al., 2017; Yip et al., 2005). The Water SARS-CoV-2 RT-PCR Test (IDEXX Laboratories, Inc.) builds on this concept by integrating two independent amplification reactions and their respective, identically labelled probes into a “single-channel multiplex” assay. The test uses primer and probe sequences from the U.S. CDC for detection of N1 and N2 (Lu et al., 2020). The output of the test is a cumulative signal from amplification and detection of both N1 and N2 target regions and the test does not quantify each target individually.

The Water SARS-CoV-2 RT-PCR Test has been used successfully for quantitative wastewater surveillance since the beginning of the pandemic (Brooks et al., 2021, 2023; Galani et al., 2022; Hopkins et al., 2023). This design has also been effective for clinical testing using a related product with EUA approval (Mascuch et al., 2020; United States Food and Drug Administration, 2020). In this report, the single-channel multiplex design used in the Water SARS-CoV-2 RT-PCR Test was examined and found to be resistant to mutations while retaining similar precision, accuracy, and reliability to the individual N1 and N2 reactions described by CDC.

## Materials & Methods

RT-qPCR was performed using the Water SARS-CoV-2 RT-PCR Test (IDEXX laboratories, Inc., 99-0015314) according to the manufacturer’s instructions. The kit provides reverse transcriptase, DNA polymerase, and a reaction buffer with all necessary components in a premixed, ready-to-use format. Reactions were assembled by combining 10 μL of RNA Master Mix, 10 μL of a primer and probe mixture (see below), and 5 μL of nucleic-acid template in a 96-well plate (Thermo Fisher Scientific, 4346906 or AB0800). Plates were sealed with film (Thermo Fisher Scientific, 4360954 or AB1170), centrifuged briefly at low speed to settle the contents and incubated in either a QuantStudio 5 (Applied Biosystems) or an AriaMx (Agilent) PCR system using the following thermocycling program: 15 min at 50°C for reverse transcription, 1 min at 95°C for denaturation and enzyme activation, followed by 45 cycles of amplification using 15 s at 95°C and 30 s at 60°C. This cycling program is standardized across different RT-qPCR tests developed by IDEXX and differs from the conditions often used for N1 and N2 amplification primarily by using a higher extension temperature (60°C). Reaction signal was detected on the FAM channel relative to a passive reference dye (ROX) included in the reaction mix and results were analyzed using the software provided by the respective manufacturer. One or more no-template controls (molecular grade water) and positive template controls (SARS-CoV-2 “PC” synthetic template; IDEXX laboratories, Inc.,) were included on each PCR plate. Quantification cycle (C_q_) results were obtained for each reaction after setting the threshold fluorescence manually at a level above background where all amplification reactions were exponential when viewed on a logarithmic plot. Within each PCR system, threshold fluorescence levels were set consistently across experiments. The data shown in Figures 1 and 2 and Tables 3, 4, and 5 were produced with the QuantStudio 5 system using a threshold value of 0.1. The data shown in Figure 3 and Table 6 was produced with the AriaMx system using a threshold value of 0.02.

**Table 1.**
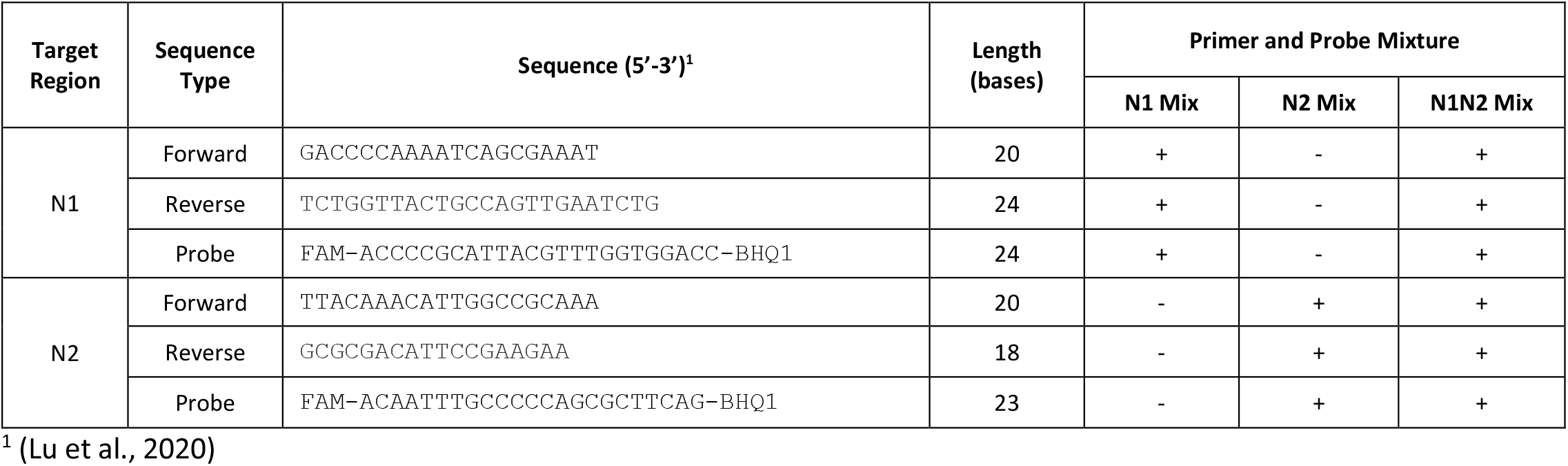
Combinations of Primers and Probes used for detection of SARS-CoV-2 by RT-qPCR.

**Table 2.**
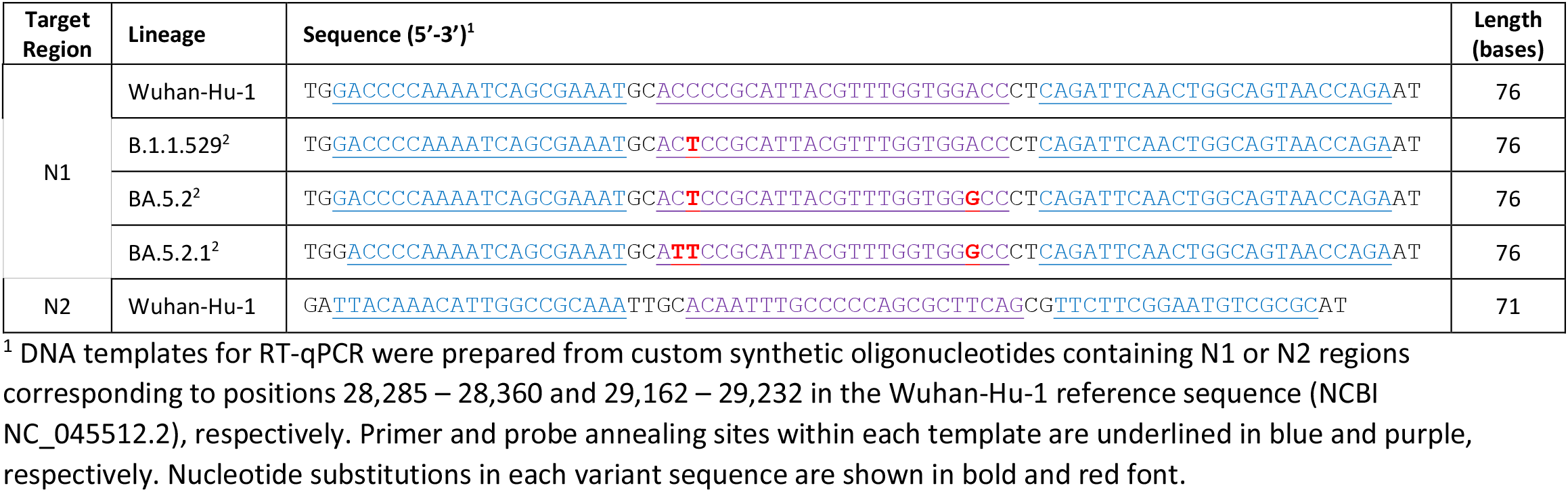
Synthetic SARS-CoV-2 DNA template sequences used for RT-qPCR.

**Table 3:** Quantitative comparison of different primer and probe combinations (N1 Mix, N2 Mix, and N1N2 Mix) for detection of single-stranded RNA containing the N gene.

| Template concentration (copies per reaction)<sup>1</sup> | N1 Mix<sup>2</sup> |  | N2 Mix<sup>2</sup> |  | N1N2 Mix<sup>2</sup> |  | Mean C<sub>q</sub> Difference<sup>3</sup> |  |  |
| --- | --- | --- | --- | --- | --- | --- | --- | --- | --- |
|  | C<sub>q</sub> | Mean C<sub>q</sub> (SD) | C<sub>q</sub> | Mean C<sub>q</sub> (SD) | C<sub>q</sub> | Mean C<sub>q</sub> (SD) | N1 Mix - N2 Mix | N1 Mix - N1N2 Mix | N2 Mix - N1N2 Mix |
| 100 | 30.8 | 31.1 (0.19) | 30.7 | 30.8 (0.08) | 29.7 | 29.8 (0.07) | 0.28 | 1.29 | 1.01 |
|  | 31.1 |  | 30.7 |  | 29.8 |  |  |  |  |
|  | 31.0 |  | 30.8 |  | 29.8 |  |  |  |  |
|  | 31.4 |  | 30.8 |  | 29.6 |  |  |  |  |
|  | 31.1 |  | 30.9 |  | 29.9 |  |  |  |  |
|  | 31.0 |  | 30.7 |  | 29.7 |  |  |  |  |
| 1,000 | 27.8 | 27.9 (0.08) | 27.4 | 27.5 (0.09) | 26.5 | 26.5 (0.07) | 0.32 | 1.34 | 1.02 |
|  | 27.8 |  | 27.5 |  | 26.4 |  |  |  |  |
|  | 27.8 |  | 27.5 |  | 26.5 |  |  |  |  |
|  | 27.9 |  | 27.5 |  | 26.6 |  |  |  |  |
|  | 28.0 |  | 27.6 |  | 26.6 |  |  |  |  |
|  | 27.8 |  | 27.6 |  | 26.5 |  |  |  |  |
| 10,000 | 24.4 | 24.5 (0.11) | 24.1 | 24.2 (0.09) | 23.0 | 23.2 (0.12) | 0.36 | 1.36 | 1.00 |
|  | 24.4 |  | 24.1 |  | 23.2 |  |  |  |  |
|  | 24.5 |  | 24.2 |  | 23.2 |  |  |  |  |
|  | 24.6 |  | 24.2 |  | 23.2 |  |  |  |  |
|  | 24.6 |  | 24.3 |  | 23.2 |  |  |  |  |
|  | 24.7 |  | 24.3 |  | 23.3 |  |  |  |  |

<sup>1</sup> RNA template added to each reaction contained wild-type N1 and wild-type N2 target regions. Template dilutions were prepared from certified RNA reference material (Quantitative Synthetic SARS-CoV-2 RNA; ATCC VR-3276SD).
<sup>2</sup>Primers and probes used in each reaction mixture shown in Table 1. Differences in detection resulted from the presence or absence of primers and probes to detect the N1 and N2 target regions. For each group of technical replicates, the mean C<sub>q</sub> and standard deviation (in parentheses) is shown.
<sup>3</sup>The difference between mean C<sub>q</sub> results was determined for each pair of reactions containing different primer and probe mixtures. Mean C<sub>q</sub> differences across all template concentrations were as follows: N1 Mix - N2 Mix: 0.32, N1 Mix - N1N2 Mix: 1.33, N2 Mix - N1N2 Mix: 1.01.

**Table 4:**
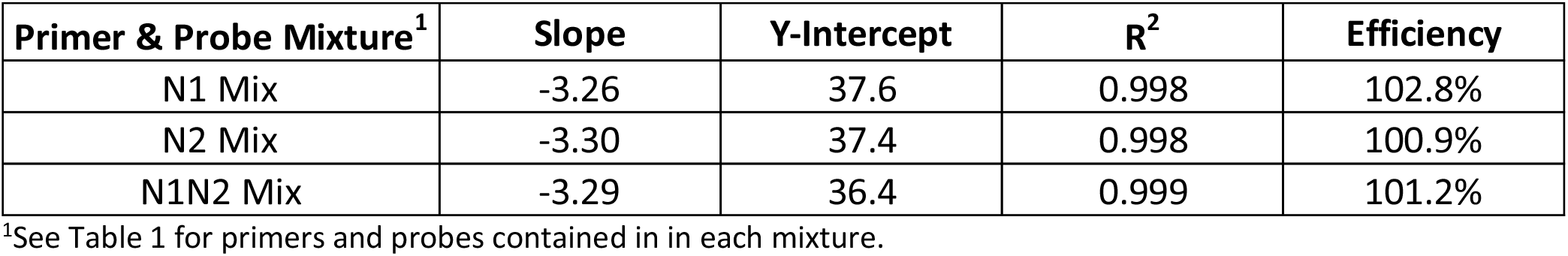
Standard curve quality parameters obtained using N1 Mix, N2 Mix, and N1N2 Mix to detect single-stranded RNA containing the N gene.

| Primer & Probe Mixture<sup>1</sup> | Slope | Y-Intercept | R<sup>2</sup> | Efficiency |
| --- | --- | --- | --- | --- |
| N1 Mix | -3.26 | 37.6 | 0.998 | 102.8% |
| N2 Mix | -3.30 | 37.4 | 0.998 | 100.9% |
| N1N2 Mix | -3.29 | 36.4 | 0.999 | 101.2% |

<sup>1</sup>See Table 1 for primers and probes contained in in each mixture.

**Table 5:** Detection of wild-type N1 or wild-type N2 DNA templates, added individually or in combination, using N1N2 Mix.

| Template concentration (copies per reaction)<sup>1</sup> | N1 template<sup>2</sup> |  | N2 template<sup>2</sup> |  | N1 & N2 template<sup>2</sup> |  | Mean C<sub>q</sub> Difference<sup>3</sup> |  |  |
| --- | --- | --- | --- | --- | --- | --- | --- | --- | --- |
|  | C<sub>q</sub> | Mean C<sub>q</sub> (SD) | C<sub>q</sub> | Mean C<sub>q</sub> (SD) | C<sub>q</sub> | Mean C<sub>q</sub> (SD) | N1 template - N2 template | N1 template - N1 & N2 template | N2 template - N1 & N2 template |
| 100 | 31.7 | 31.9 (0.40) | 32.5 | 32.6 (0.31) | 30.7 | 30.9 (0.21) | -0.63 | 1.06 | 1.69 |
|  | 32.7 |  | 33.0 |  | 30.7 |  |  |  |  |
|  | 31.8 |  | 32.5 |  | 30.8 |  |  |  |  |
|  | 32.0 |  | 32.8 |  | 30.9 |  |  |  |  |
|  | 32.0 |  | 32.4 |  | 31.2 |  |  |  |  |
|  | 32.2 |  | 32.5 |  | 31.0 |  |  |  |  |
|  | 31.7 |  | 32.0 |  | 31.1 |  |  |  |  |
|  | 31.4 |  | 32.8 |  | 30.6 |  |  |  |  |
| 1,000 | 28.1 | 28.4 (0.18) | 28.8 | 29.1 (0.20) | 27.2 | 27.5 (0.17) | -0.64 | 0.93 | 1.57 |
|  | 28.5 |  | 29.0 |  | 27.4 |  |  |  |  |
|  | 28.4 |  | 29.0 |  | 27.4 |  |  |  |  |
|  | 28.7 |  | 29.1 |  | 27.7 |  |  |  |  |
|  | 28.5 |  | 29.1 |  | 27.6 |  |  |  |  |
|  | 28.6 |  | 29.2 |  | 27.7 |  |  |  |  |
|  | 28.4 |  | 29.5 |  | 27.7 |  |  |  |  |
|  | 28.4 |  | 28.9 |  | 27.4 |  |  |  |  |
| 10,000 | 24.8 | 24.9 (0.12) | 25.4 | 25.6 (0.11) | 23.8 | 24.1 (0.14) | -0.72 | 0.80 | 1.53 |
|  | 24.7 |  | 25.6 |  | 24.1 |  |  |  |  |
|  | 24.9 |  | 25.6 |  | 24.2 |  |  |  |  |
|  | 24.9 |  | 25.6 |  | 24.3 |  |  |  |  |
|  | 25.0 |  | 25.8 |  | 24.0 |  |  |  |  |
|  | 25.0 |  | 25.7 |  | 24.1 |  |  |  |  |
|  | 25.0 |  | 25.7 |  | 24.1 |  |  |  |  |
|  | 24.9 |  | 25.5 |  | 24.1 |  |  |  |  |

<sup>1</sup>DNA templates added to each reaction contained wild-type N1 and wild-type N2 target regions (Table 2). Each template (N1 or N2) was added at the indicated concentration.
<sup>2</sup>All reactions were performed using N1N2 Mix containing primers and probes for detection of both N1 and N2 regions. Differences in detection resulted from the presence or absence of N1 and N2 template regions. For each group of technical replicates, the mean C<sub>q</sub> and standard deviation (in parentheses) was determined. Results for each reaction were significantly different (p<0.05).
<sup>3</sup>The difference between mean C<sub>q</sub> results was determined for each pair of reactions containing different template combinations. Mean C<sub>q</sub> differences across all three template concentrations were as follows: N1 template - N2 template: -0.66, N1 template – N1 & N2 templates: 0.93, N2 template – N1 & N2 templates: 1.59.

**Table 6:**
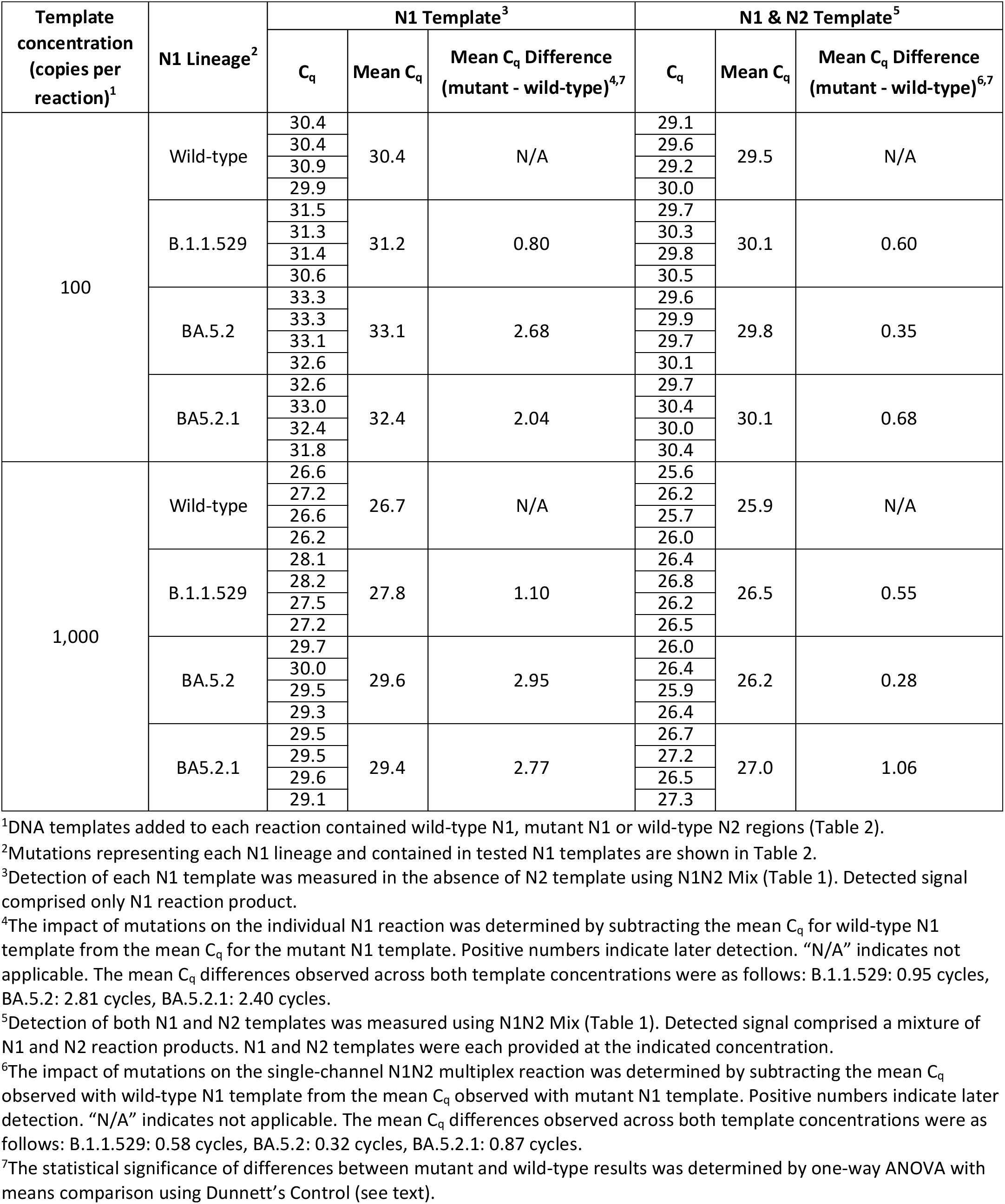
Quantitative detection of wild-type and mutant N1 template sequences using a single-channel multiplex reaction.

**Figure 1:**
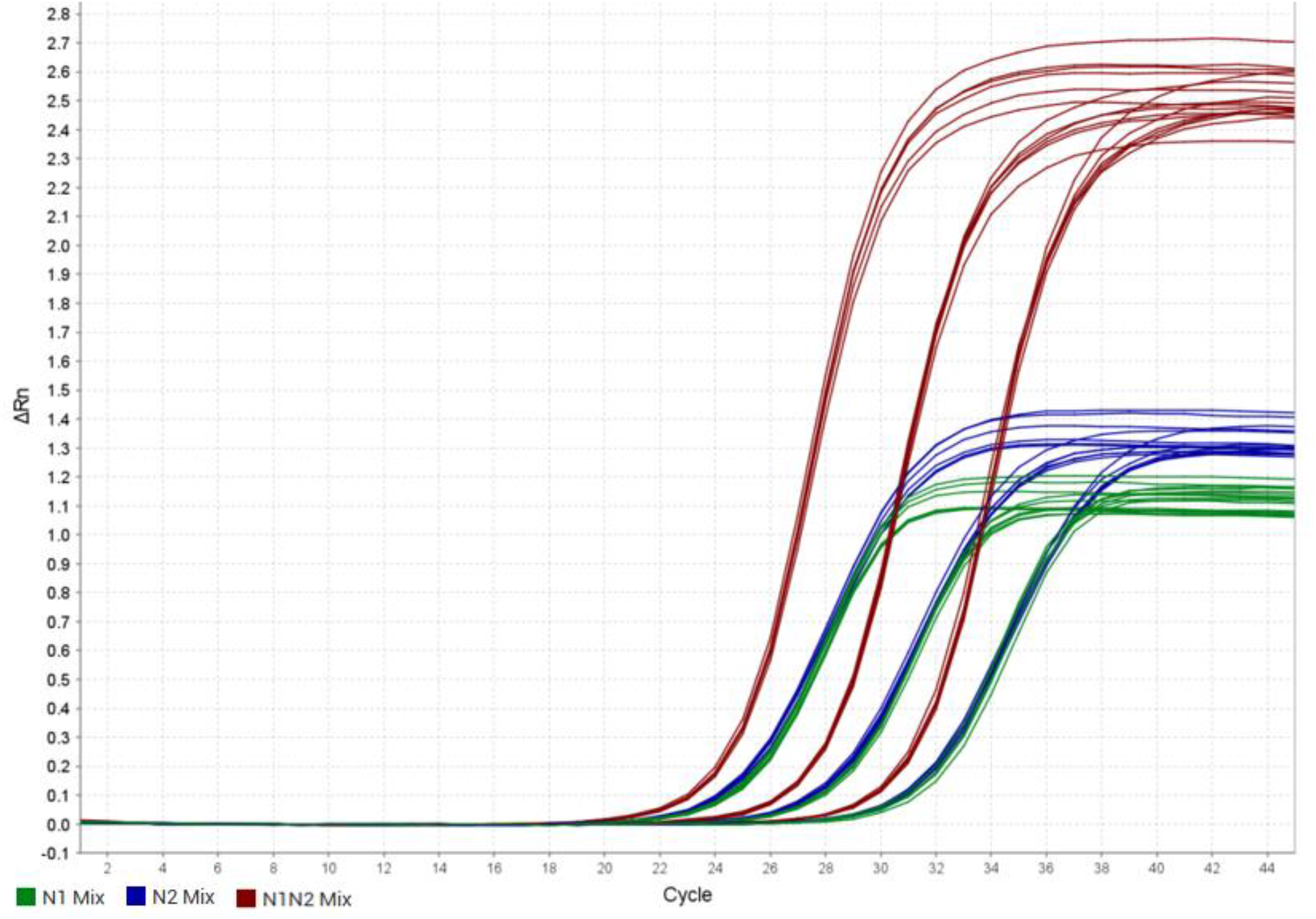
Detection of single-stranded RNA containing N1 and N2 regions at 100, 1,000, and 10,000 copies per reaction. Primer and probe combinations tested included N1 Mix (green) and N2 Mix (blue) and N1N2 Mix (red) (Table 1). All reactions used probes labelled with FAM fluorophore.

**Figure 2:**
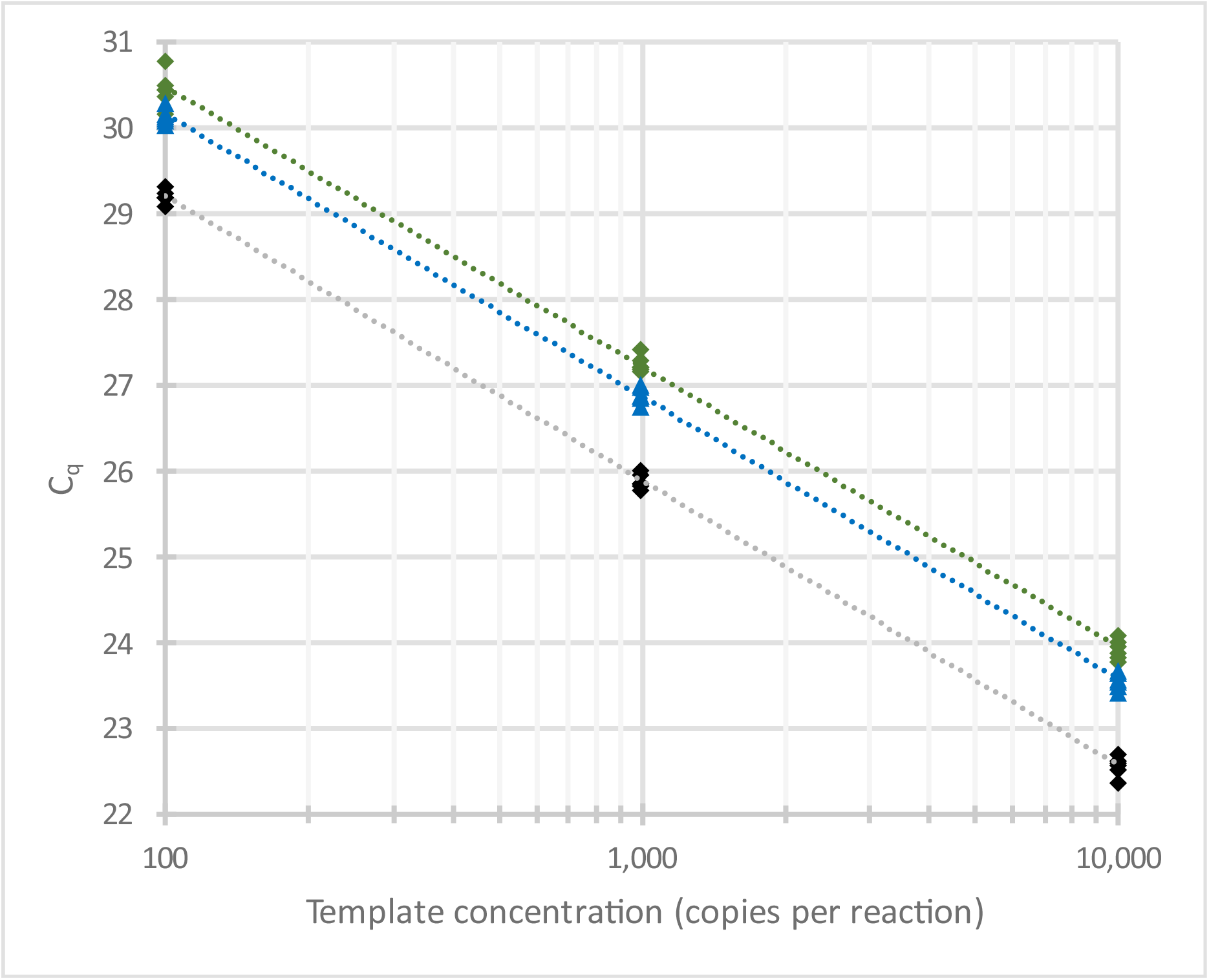
Standard curves produced using N1 Mix (green), N2 Mix (blue), and N1N2 Mix (black) to detect single-stranded RNA containing the N gene at 100, 1,000, and 10,000 copies per reaction.

**Figure 3:**
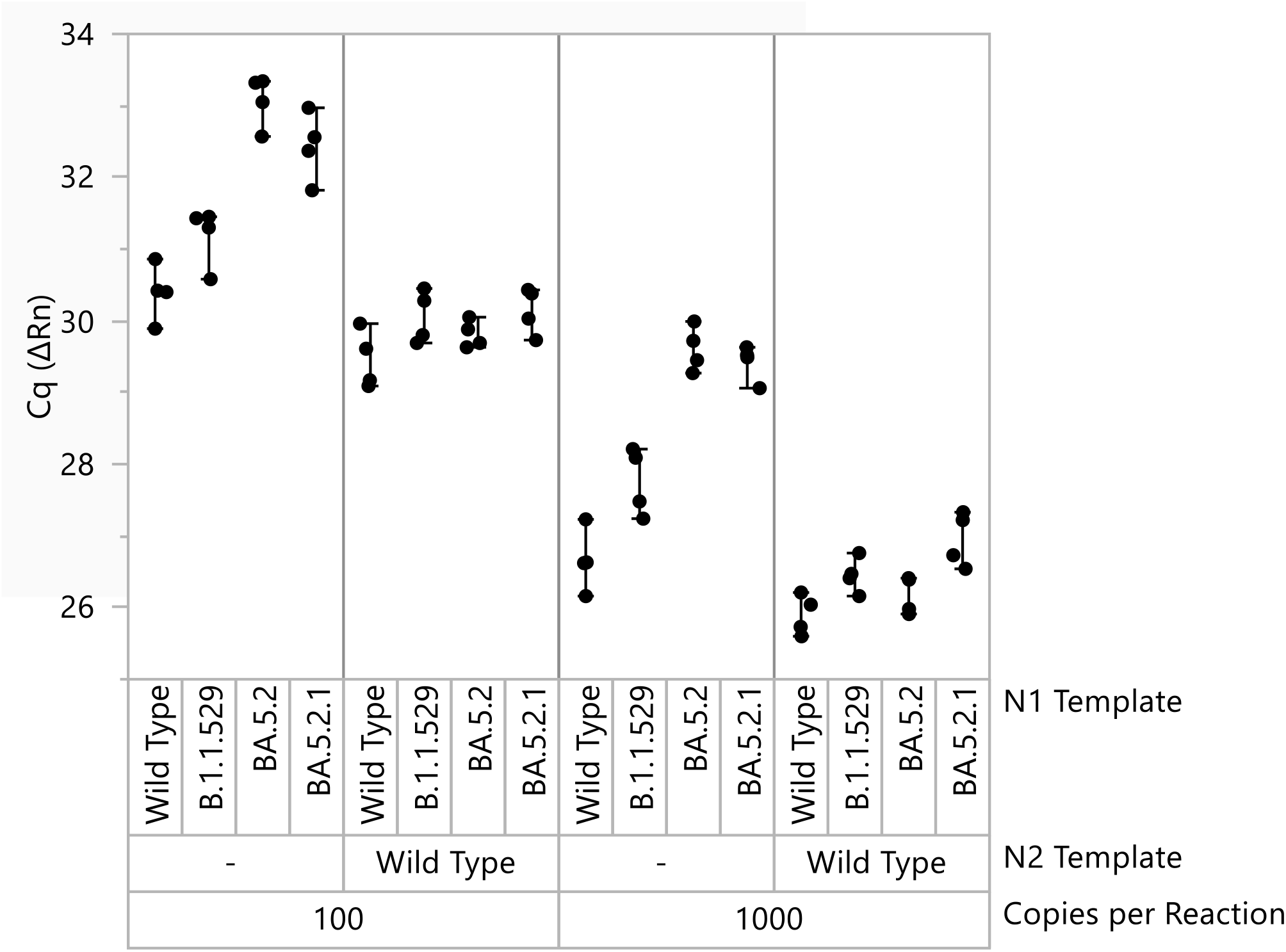
Impact of mutations from three different Omicron lineages on the individual N1 reaction and the single-channel N1N2 multiplex reaction. All reactions were performed using N1N2 Mix (Table 1). Reaction signal varied according to the presence of N1 and N2 template. Separate reactions were performed in which each N1 template sequence was tested in the presence and absence of wild-type N2 template. When only N1 template was provided, fluorescent signal was produced solely from N1 amplification and detection corresponding to individual N1 detection. When both N1 and N2 template were provided, fluorescent signal was produced from simultaneous amplification and detection of N1 and N2 corresponding to the N1N2 single-channel multiplex reaction.

### Primers and probes

Custom derivatives of the primer and probe mixture contained in the Water SARS-CoV-2 RT-PCR Test were used in this study. The kit provides a ready-to-use mixture of the primers and probes described by the US CDC for detection of N1, N2, and Human RNaseP (Lu et al., 2020). To perform the full reaction as used in the IDEXX Water SARS-CoV-2 RT-PCR Test, primers and probes for all three reactions (N1, N2, and Internal Control) were combined in a single mixture (N1N2 Mix) at final concentrations similar to the recommendations from Lu et al. Custom reactions were also prepared excluding either the N2 reaction (N1 Mix) or N1 reaction (N2 Mix). A summary of the different primer and probe combinations used in this study is provided in Table 1. Probes for both N1 and N2 were identically labelled at the 5′ end with 6-carboxy-fluorescein (FAM) and at the 3′ end with Black Hole Quencher 1. All primers and probes were prepared and diluted in 10 mM Tris-HCl pH 8.0 with 0.1 mM EDTA (low EDTA TE Buffer, Sigma T2694 or Invitrogen 12090-015) supplemented with approximately 0.05 mg/mL Poly A (Sigma 10108626001). The statistical significance of differences in variance between each combination of primers and probes was evaluated with Levene’s and Bartlett’s tests. Statistical analyses were conducted using JMP software (JMP Statistical Discovery).

### Template

RNA template was prepared from a commercial product (Quantitative Synthetic SARS-CoV-2 RNA; ATCC VR-3276SD). Based on the initial concentration provided by the manufacturer, dilutions were prepared in TE buffer with Poly A to provide 100, 1,000, or 10,000 copies per reaction. DNA templates containing wild-type N1 or N2, or mutant variants of N1, were synthesized as custom single-stranded oligos (Thermo Fisher Scientific). A summary of the DNA template sequences used in this study is shown in Table 2. Each synthetic was dissolved in TE with Poly A and diluted to a target final concentration of 100, 1,000, or 10,000 copies per reaction assuming complete resuspension of the molar synthesis yield provided by the manufacturer. The concentration of each template was verified through determination of the most probable number (MPN) from a proportion of positive and negative results (Hurley & Roscoe, 1983). To obtain MPN estimates, each resuspended DNA template was diluted to approximately one copy per reaction in TE with Poly A. A range of 28 to 30 replicate PCR reactions was analyzed for the presence or absence of product signal. DNA templates containing the corresponding mutations found in the N1 region of specific Omicron variants (BA.1, BA.5.2, and BA.5.2.1) were synthesized and prepared using the same method. The statistical significance of mean C_q_ results obtained with each mutant template was evaluated by one-way ANOVA with comparison of means by Dunnett’s Method using wild-type template results as the control.

## Results

Three sets of experiments were performed to characterize the N1N2 single-channel multiplex design used in the IDEXX SARS-COV-2 Test. First, qualitative and quantitative performance of the N1N2 single-channel multiplex, including standard curve properties, was compared with results obtained using individual N1 and N2 reactions. Testing was performed using different combinations of primers and probes (Table 1) and all reactions included a single-stranded RNA template containing both wild-type N1 and N2 regions. Second, the N1N2 single-channel multiplex was tested with different combinations of DNA templates. Testing was performed by providing synthetic sequences containing either N1, N2, or a mixture of the N1 and N2 regions at equal concentration (Table 2). Third, the impact of real-world mutations in the N1 probe annealing region was examined. The relative performance of the N1N2 single-channel multiplex and the individual N1 and N2 reactions was compared using three different mutant N1 template sequences found in recent Omicron lineages (Table 2).

### Characterization of N1N2 Single-Channel Multiplex

The N1N2 single-channel multiplex and individual N1 and N2 reactions were tested with three concentrations of RNA template containing the N gene. Results for all conditions are shown in Figure 1, and a summary of C_q_ values obtained for all technical replicates is provided in Table 3. The results show that the individual N1 and N2 reactions, when tested separately, produce very similar amplification curves. The N1N2 single-channel multiplex reaction, which detects a composite signal from the N1 and N2 reactions using one fluorophore (FAM), produced a stronger fluorescence that was detected earlier in the reaction, with no significant change in background affecting threshold values compared to the individual N1 and N2 assays. Closer inspection showed that fluorescent signal from the N1N2 multiplex was an additive combination of the individual N1 and N2 reactions. Consistent with these observations, the mean C_q_ values obtained across all template concentrations showed the individual N2 reaction produced virtually the same result as the individual N1 reaction with a difference of only 0.32 cycles. The combined N1N2 reaction showed earlier detection by 1.33 and 1.01 cycles when compared to the individual N1 and N2 reactions, respectively. The intra-assay repeatability of the N1N2 multiplex was not significantly different (p>0.05) from the individual N1 and N2 reactions and all three reactions showed high precision, with low C_q_ standard deviations ranging from 0.07 to 0.19 across all template concentrations (Table 3). Collectively these results demonstrated the N1N2 multiplex reaction to behave consistently and predictably.

To assess quantitative performance of each primer and probe combination, the data presented in Table 3 were used to generate three-point standard curves (Figure 2 and Table 4). The use of six technical replicates at each template concentration allowed reliable comparisons to be made. All three primer and probe mixtures produced consistent standard curves with high R^2^ values >0.997 and efficiency values ranging from 100 to 103%. These compare favorably to MIQE guidance values of >0.98 R^2^ and 90 to 110% efficiency (Bustin et al., 2009). Reactions containing the N1 Mix or N2 Mix produced very similar results. Results with the N1N2 Mix showed no significant difference in slope, R^2^, or efficiency relative to N1 Mix or N2 Mix. The N1N2 Mix produced a y-intercept shift of 1.1 cycles as expected due to the observed bias in C_q_ results between the reactions (Table 3).

The obtained standard curves allowed the N1N2 reaction to produce the same quantitative result as the individual N1 and N2 reactions. For example, the concentration of the 1,000-copy dilution was back-calculated from its mean C_q_, as if it was an unknown sample. Results obtained with the N1 Mix and N2 Mix produced respective concentrations of 971 and 972 copies per reaction using the corresponding standard curve for each reaction. Similarly, results obtained with the N1N2 Mix produced a virtually identical result of 980 copies per reaction. Analogous results were obtained across the entire range of the standard curve, with the N1N2 reaction showing a maximum difference in calculated result from the N1 or N2 assays of 1.0%. These data demonstrate that the N1N2 single-channel multiplex reaction provided similar quantitative performance with no significant difference in accuracy or precision from individual N1 or N2 reactions.

### Results with N1 and N2 DNA templates

The combined N1N2 reaction was further characterized in a second complementary approach where the N1N2 Mix was tested with different templates. Experiments were performed in which single-stranded DNA oligos, containing either the wild-type N1 or N2 target region, were added individually or in combination as reaction template. Tested DNA oligos are shown in Table 2.

When tested individually with N1N2 Mix, similar C_q_ values were observed for the wild-type N1 and N2 templates, with N1 detected slightly earlier than N2 by an average of 0.66 cycles across all three template concentrations (Table 5). When the wild-type N1 and N2 templates were combined, the N1N2 Mix provided earlier detection than was observed with the individual N1 (0.93 cycles) or N2 (1.59 cycles) templates. Similar precision was observed with each template (Table 3) and intra-assay repeatability across the three templates was not significantly different (p>0.05). These results were generally consistent with the findings above using RNA where (1) detection of the individual target regions using N1 Mix and N2 Mix produced similar C_q_ results, and (2) simultaneous detection of the N1 and N2 regions using N1N2 Mix showed earlier detection than either individual reaction (Table 3).

The consistent results obtained from both sets of experiments (Tables 3 and 5) suggest that potential artifacts of these experimental approaches were unlikely to have had a large effect. The most likely source of error was in the second experiment where differences in the concentration of the N1 and N2 templates could contribute to the observed differences in C_q_ (Table 5). To evaluate this possibility the concentration of each template preparation was estimated using a most probable number (MPN) determined from a proportion of positive and negative results obtained when each template was diluted to approximately one copy per reaction. For the wild-type N1 template, 13 of 28 replicate reactions showed positive results, producing an MPN of 0.62 copies per reaction (95% confidence interval: 0.36 to 1.1). Similarly, for the wild-type N2 template, 15 of 30 replicate reactions were positive, producing an MPN of 0.69 copies per reaction (95% confidence interval: 0.41 to 1.2).

These results confirmed the relative concentrations of each template were not meaningfully different and both N1 and N2 were present at the approximate concentrations indicated in Table 5.

### Performance with mutant templates

Collectively the results above show the primers and probes for detection of N1 and N2 perform similarly and provide a predictable, additive result when combined on a single fluorescent channel. Importantly, the maximum difference observed between the single-channel N1N2 multiplex reaction and individual N1 or N2 reactions was relatively small and only slightly larger than one cycle (range 0.93 to 1.59; Tables 3 and 5). It is expected that the single-channel N1N2 multiplex reaction would show a maximum C_q_ increase of similar magnitude if either the N1 or N2 reaction was completely disrupted by any another mechanism, such as a mutation in a primer or probe annealing site. These results predict that the N1N2 multiplex reaction will be less sensitive to mutation than either the individual N1 or N2 reactions.

To test this hypothesis, the impact of real-world mutations in the N1 target region was examined with the N1N2 multiplex reaction. Analysis of SARS-CoV-2 sequences obtained from the GISAID database (GISAID, 2020) throughout the pandemic initially showed the N1 and N2 annealing regions to be affected only by rare and sporadic mutations that were not enriched in the circulating population (data not shown). Sequences analyzed between December 2021 and March 2023 related to the dominant Omicron variant lineages showed the first evidence of a prevalent mutation, defined by the authors as occurring in >5% of the sequence population, affecting the probe annealing site in N1 (data not shown). Later Omicron variants showed additional prevalent mutations also affecting the N1 probe annealing site. Here, three combinations of these nucleic acid substitutions in N1 were selected for study, representing those found in Omicron variants B.1.1.529, BA.5.2, and BA.5.2.1 (Table 2).

Testing was performed using two conditions in a similar experimental approach to that described above. First, the impact of each mutant sequence on N1 detection was measured in the absence of N2 template. Second, the impact of each mutant sequence on combined detection of N1 and N2 using the single-channel multiplex reaction was measured when both N1 and N2 templates were added to the reaction. The second condition mimics the natural detection of SARS-CoV-2 where detection of a single viral genome containing intact N gene is accomplished by simultaneous amplification of both the N1 and N2 regions. A summary of the results is shown in Figure 3 and Table 6.

Results with the individual N1 templates showed that each mutation reduced the effectiveness of detection, with the B.1.1.529, BA.5.2, and BA.5.2.1 sequences showing mean C_q_ increases compared to wild-type of 0.95, 2.81, and 2.40 cycles, respectively, across both template concentrations tested. The C_q_ results for each mutant obtained with the individual N1 reaction were found to be significantly different (p<0.05) from the wild-type control in all conditions tested. These relative differences were consistent with the expectation that the double and triple mutations contained in BA.5.2 and BA.5.2.1 templates, which reside at both ends of the N1 probe annealing region, would be more disruptive than the single mutation found in B.1.1.529. Reduced detection of BA.5.2 and BA.5.2.1 produced a 5.3-fold to 7.0-fold relative underestimation of the actual template concentration, representing a significant reduction in accuracy for the individual N1 reaction.

In contrast, the single-channel N1N2 multiplex reaction was more resistant to mutation, with the B.1.1.529, BA.5.2, and BA.5.2.1 sequences showing average increases of 0.58, 0.32, and 0.87 cycles, respectively, across both template concentrations tested. These results showed that the N2 reaction compensated almost entirely for the defects in N1 reaction caused by mutation and produced a much smaller range of underestimation of only 1.3-fold to 1.8-fold (Table 5). The statistical significance of these results varied among the conditions tested. At the 100-copy level, all three mutant templates were not significantly different (p>0.05) from the wild-type control when tested with the N1N2 multiplex reaction. At the 1000-copy level, only the BA.5.2 mutant was not significantly different (p>0.05) from the wild-type control. Taken together, the effect of each mutation on the N1N2 multiplex was consistently smaller than the corresponding differences observed with the individual N1 assay, and the impacts of such mutations on the N1N2 assay are unlikely to have a material impact on wastewater surveillance trending (see Discussion). Similar results were obtained in a second set of experiments using independently obtained and prepared synthetic materials (data not shown).

Although this study has specifically examined mutations in the N1 probe annealing region due to their importance for surveillance of Omicron lineages, it is expected that the single-channel N1N2 multiplex reaction will be similarly less sensitive to mutations occurring in either the N1 or N2 annealing regions, including changes affecting either primer or probe annealing regions. This is consistent with the results above showing a maximum shift of approximately one cycle when either individual reaction was completely disrupted (Table 5).

## Discussion

Previous work by Nagy et al. (2017) and Yip et al. (2005) showed the advantages of using identically labelled probes to increase target inclusivity and reduce false negative results caused by mutations in probe binding regions. In this study it was shown that similar benefits can be achieved using a broader approach in which two independent amplification reactions that each use an identically labelled probe are combined. An important benefit of this single-channel multiplex approach is that the entirety of each assay design, including both primer and probe annealing regions, becomes more resistant to mutations.

Our results with the single-channel N1N2 multiplex assay demonstrated similar performance characteristics to those reported by Nagy et al. and Yip et al., including stronger fluorescence intensity, earlier C_q_ results, high precision in intra-assay technical replicates, accurate standard curve performance, and improved detection of mutant sequences. Our results also showed that the N1N2 assay achieved these benefits without altering the performance characteristics of the individual N1 and N2 reactions. Taken together, these results show that the N1N2 multiplex assay reduced the impact of real-world mutations in the N1 region while retaining consistent and accurate quantification of SARS-CoV-2 with equivalent performance to the individual N1 or N2 assays.

Although the N1 and N2 reactions were designed in conserved regions, prevalent sequence changes have recently been observed in the N1 region of Omicron variant lineages and these mutations were found to significantly reduce the accuracy of an individual N1 reaction. In real-world surveillance of wastewater containing such Omicron variants, a laboratory using individual N1 and N2 assays would likely observe a growing divergence in results between the two assays caused by these mutations, with a concomitant increase in uncertainty of results interpretation. In contrast, a laboratory using the N1N2 multiplex assay is expected to observe only small changes in trending accuracy as such mutations emerge and spread through a monitored community. The average relative impact of the Omicron mutations on the N1N2 multiplex was a C_q_ shift of ≤0.87 cycles (Table 6). Furthermore, in the extreme situation where detection of either N1 or N2 was completely absent, the maximum impact on the N1N2 multiplex was a C_q_ shift of 1.33 cycles (Tables 3 and 5). This is only slightly larger in magnitude than an acceptable margin of error for RT-qPCR recently recommended for wastewater surveillance (<0.5 standard deviation for technical triplicate measurements) (Chik et al., 2022). When considered in the context of the overall wastewater testing process, the effect of these mutations on the N1N2 multiplex assay was insignificant compared to other, larger sources of variation existing in upstream wastewater concentration and handling steps (Pecson et al., 2021).

The single-channel multiplex N1N2 assay described here provides a reliable and advantageous approach for RT-qPCR surveillance with target regions that are susceptible to mutation. In comparison, the larger impact of mutations on individual N1 and N2 assays can reduce confidence in results interpretation and increase the need for auxiliary information from complementary monitoring strategies, such as sequencing, to assess the impact of emerging mutations. Although sequencing is inherently valuable and should be used regardless of RT-qPCR assay design, the interpretation of results from the single-channel N1N2 multiplex assay is less dependent on sequencing data. This allows public-health decisions to be made more quickly and confidently from trending data produced using the N1N2 multiplex test. The multiplex assay also allows a simpler testing process that requires fewer RT-qPCR reactions and can reduce the potential for technical errors. Collectively, this study showed that the single-channel N1N2 multiplex assay can be used in place of individual N1 and N2 reactions to increase the reliability and accuracy of wastewater surveillance trending data for the evolving SARS-CoV-2 target. Given these advantages, it is expected that a similar single-channel approach will be beneficial in other surveillance programs that monitor nucleic acid targets susceptible to mutation.

## Data Availability

All data produced in the present study are available upon reasonable request to the authors

